# The role of remdesivir in South Africa: preventing COVID-19 deaths through increasing ICU capacity

**DOI:** 10.1101/2020.06.10.20127084

**Authors:** Brooke E Nichols, Lise Jamieson, Sabrina RC Zhang, Sheetal Silal, Juliet R.C. Pulliam, Ian Sanne, Gesine Meyer-Rath

## Abstract

Countries such as South Africa have limited intensive care unit (ICU) capacity to handle the expected number of COVID-19 patients requiring ICU care. Remdesivir can prevent deaths in countries such as South Africa by decreasing the number of days people spend in ICU, therefore freeing up ICU bed capacity.

## MANUSCRIPT

The COVID-19 pandemic has already infected millions of people across the globe, with more than 400,000 COVID-related deaths reported worldwide.(1) Case numbers are growing rapidly in low-and middle-income countries, overwhelming health systems. In South Africa, the combined intensive care unit (ICU) capacity of the public and private health sectors combined was estimated to be 3,340 before the COVID-19 pandemic took hold. As of the beginning of June 2020, ICU beds in the Western Cape, current the worst affected of South Africa’s nine provinces, were already at capacity,(2) and other provinces’ capacity is projected to be overwhelmed in the coming weeks. ICU capacity is expected to be breached, depending on the provinces, for between 3-6 months between May-December 2020 by a factor of 6-10.(3)

There are currently no curative therapies available for those with advanced COVID-19 disease, and mortality rates among those requiring ICU care are around 50%.(4, 5) Recently, while showing limited direct impact on morbidity and mortality, remdesivir has been shown to reduce the total amount of time in ICU required per patient, from 15 days on average (95% confidence interval [CI] 13-19 days), down to 11 days on average (95%CI 9-12 days).(6) Given the limited ICU capacity in South Africa, we sought to determine the impact that remdesivir could have in preventing deaths through decreasing the amount of time people spend in ICU, and, as a result, increasing ICU bed turnover.

We first determined the number of people requiring ICU admission by province by month as projected by the South African National COVID-19 Epidemiology model (NCEM) between May and December 2020, and the number of all currently available ICU beds by province.(3) We assumed that remdesivir would be used for all ICU patients in South Africa during peak months where ICU capacity is expected to be breached, i.e. June-September 2020. We used 10,000 Monte Carlo simulations to sample uniformly within the 95% confidence interval (CI) of recovery time for patients treated with and without remdesivir using the results of a recently published remdesivir clinical trial. We assumed that the mortality rate of those requiring ICU care and receiving ICU care was 50%,(4, 5) and varied the death rate of those requiring ICU care but not receiving ICU care between 85-100%. We projected outcomes for two scenarios defined by optimistic and pessimistic assumptions regarding the impact of the country’s mitigation strategies since April 2020 on transmission, and report the 2.5^th^-97.5^th^ percentiles of the Monte Carlo simulations.

In South Africa, there are an estimated 3,450 ICU beds available for COVID patients. ICU capacity is expected to be breached, depending on the province, for between three and six months between May and December 2020. Without remdesivir, there is expected to be a total of 23,416-31,269 people that will occupy an ICU bed during peak months where ICU is expected to be breached (**Table**). With remdesivir, that number is expected to grow by 51-54% to 36,291-47,827 treated in the ICU by December 2020. By giving remdesivir to all patients in ICU during peak months (all 36,291-47,827 patients), 3,295-6,814 deaths will be averted if assuming an 85% death rate amongst those not receiving ICU care but require ICU care, and 4,707-9,734 deaths averted if assuming a 100% death rate amongst those not receiving ICU care but require ICU care. This substantial impact is mediated solely through reducing the number of ICU days required per patient, and thereby creating additional capacity to treat more patients.

This analysis has several limitations. Firstly, if the epidemic curve is altogether flatter than predicted in our model projections, then the total number of deaths averted could be higher, as the number requiring ICU care would be spread out over a greater number of months. As the epidemic matures, the composition of patient population entering the ICU may change, particularly if any rationing of resources occurs. While this may affect the point estimate of our results, it is unlikely to affect the magnitude of difference between scenarios. Importantly, while the simple mechanism of reducing the number of ICU days required will have a large impact on mortality, it can only have an impact that is only as large as it’s ICU infrastructure. For countries that have an even more constrained ICU bed capacity, such as Zambia, with just 91 ICU beds,(7) the absolute benefit will be much smaller.

To conclude, remdesivir, or any intervention that reduces ICU length of stay, can play a significant role in freeing up highly constrained ICU capacity, and therefore reduce the number of deaths caused by limited capacity. South Africa and countries with similarly constrained healthcare system capacity are expected to benefit the most and should consider remdesivir, or any other interventions that can reduce length of ICU stay, for patients in ICU with COVID-19-if prices (which are currently still under negotiation) are acceptable.

**Table.** Estimates of patients treated in ICU, and expected deaths averted due to shortened ICU stay with remdesivir treatment; Monte Carlo simulation (n=10,000) results presented with medians and 2.5^th^ and 97.5^th^ percentiles

|  | Optimistic<br>(median, 2.5<sup>th</sup> -97.5<sup>th</sup><br>percentile) | Pessimistic<br>(median, 2.5<sup>th</sup> -97.5<sup>th</sup><br>percentile) |
| --- | --- | --- |
| Total number of patients receiving ICU care during months where capacity is breached, **no remdesivir** | 25,738<br>(23,416 - 28,412) | 29,314<br>(26,694 - 32,269) |
| Total number of patients receiving ICU care during months where capacity is breached, **100% on remdesivir** | 39,068<br>(36,291 - 42,103) | 44,493<br>(41,352 - 47,827) |
| Additional patients receiving ICU care due to remdesivir during months where capacity is reached | 13,307<br>(9,414 - 17,251) | 15,164<br>(10,838 - 19,467) |
| Mortality if those additional patients receive ICU care (50%) | 6,654<br>(4,707 - 8,625) | 7,582<br>(5,419 - 9,734) |
| Mortality if those additional patients do not receive ICU care (85% mortality assumed) | 11,311<br>(8,002 - 14,663) | 12,890<br>(9,212 - 16,547) |
| Mortality if those additional patients do not receive ICU care (100% mortality assumed) | 13,307<br>(9,414 - 17,251) | 15,164<br>(10,838 - 19,467) |
| **Deaths averted (low, 85% mortality)** | **4,657**<br>**(3,295 - 6,038)** | **5,307**<br>**(3,793 - 6,814)** |
| **Deaths averted (high, 100% mortality)** | **6,654**<br>**(4,707 - 8,625)** | **7,582**<br>**(5,419 - 9,734)** |

## Data Availability

Only publicly available data were used for this analysis.

